# Cell-free telomeric DNA and integrity indices in axial spondyloarthritis: associations with somatic and mental burden — a case-control study

**DOI:** 10.64898/2026.11.15.26368092

**Authors:** Markéta Hušáková, Bhaswarupa Banerjee, Patrik Škubica, Karel Pavelka, Karolína Kuchaříková, Jan Kotrs, Ladislav Šenolt, Marta Olejárová, Pavel Mohr, Pavlína Daňková

**Affiliations:** First Faculty of Medicine, Charles University and Rheumatology Institute, Na Slupi 4, Prague, Czech Republic; Department of Anthropology and Human Genetics, Faculty of Science, Charles University, Prague, Czech Republic; Department of Public Mental Health, National Institute of Mental Health, Klecany, Czech Republic; Health Monk s.r.o., Prague, Czech Republic; Faculty of Informatics, Masaryk University, Brno, Czech Republic; Clinical Center, National Institute of Mental Health, Klecany, Czech Republic; Third Faculty of Medicine, Charles University, Prague, Czech Republic

**Keywords:** Axial spondyloarthritis, depression, anxiety, inflammation, CRP, cell-free DNA

## Abstract

**Objectives:** Depression and anxiety in axial spondyloarthritis (axSpA) are associated with higher disease activity. This study examined whether circulating cell-free nuclear DNA (cf-nDNA), mitochondrial DNA (cf-mtDNA), relative telomere length (T/S ratio), and DNA integrity index (DII) are associated with mental burden and disease characteristics in axSpA.

**Methods:** Plasma cf-nDNA, cf-mtDNA, T/S ratio, and ALU-247/ALU-115 ratio (DII) were analysed in 79 patients with axSpA and 88 age- and sex-matched healthy controls (HCs). Mental burden was assessed using the Mini-International Neuropsychiatric Interview (M.I.N.I.), Beck Depression Inventory-II, and Beck Anxiety Inventory. Clinical measures included the Bath Ankylosing Spondylitis Disease Activity Index (BASDAI), Ankylosing Spondylitis Disease Activity Score with C-reactive protein (ASDAS-CRP), and radiographic disease pattern.

**Results:** Sex differences were observed in disease phenotype and hip involvement. Affective or anxiety disorders were more frequent in axSpA vs. HC (26.6% vs. 2.3%, p<0.001); depressive symptoms were also more frequent (20.2% vs. 2.27%, p<0.001) and higher in active vs. inactive disease (30.0% vs. 3.45%, p<0.001). Compared with HCs, patients had higher cf-mtDNA and lower DII (both p<0.01), with a trend towards lower T/S ratio. Females had higher T/S ratio and DII than males, and higher cf-mtDNA overall. Lower T/S ratio was associated with affective disorders, while only CRP independently predicted depressive symptom severity.

**Conclusions:** Altered cfDNA profiles, particularly increased cf-mtDNA and reduced DII, are associated with axSpA, disease activity and mental burden, supporting their potential as biomarkers of disease-related processes.

## Introduction

Axial spondyloarthritis (axSpA) is a painful and disabling chronic inflammatory disease characterised by arthritis and enthesitis, with inflammatory processes affecting the spine and sacroiliac joints (SIJs), leading to subsequent structural damage [1]. Immune dysregulation involving both innate and adaptive immune mechanisms, particularly Th17-driven pathways, promotes musculoskeletal and extra-articular inflammatory manifestations of axSpA [2]. Inflammation affecting additional tissues and cell types further contributes to the development of multiple comorbidities, including cardiovascular diseases, infections, and increased psychological distress [3].

Patients with axSpA have been shown to have a higher risk of major depression (MDD), with a pooled relative risk (RR) of 1.51 (95% confidence interval [CI] 1.28–1.79) [4] as well as anxiety disorders (AnxD), with a hazard ratio (HR) of 1.84 (95% CI 1.37–2.49), compared with the general population [5]. These associations have been further explored at the population level in large-scale studies of axSpA. The International Map of Axial Spondyloarthritis (IMAS) study recently identified mental health burden as a global challenge in axSpA, reporting that 40.5% and 37.2% of patients experienced anxiety and depressive symptoms, respectively [6]. In axSpA, depression and anxiety have been predominantly associated with higher disease activity, while no consistent association has been observed with radiographic severity of the SIJs [7] or even treatment response [8]. Beyond the psychosocial aspects of axSpA, immune-brain interactions may represent a possible link to depression and anxiety. Specifically, elevated C-reactive protein (CRP) levels have been reported in patients with MDD [9], with inflammation being particularly associated with somatic depressive symptoms, including reduced energy, sleep disturbances, exhaustion, low mood, and psychomotor slowing [10].

Various types of extracellular DNA fragments, termed circulating cell-free (cf)DNA, are released into the bloodstream as a result of apoptosis or other types of cell death, cellular damage, and NETosis that accompany cancer, stress, trauma, and intensified immune responses, including host defence against infections, antitumour immunity, and autoimmune processes. Following cellular uptake, cfDNA engages intracellular DNA sensors, including Toll-like receptor 9, cyclic GMP–AMP synthase, and absent in melanoma 2, thereby activating downstream signalling pathways that induce pro-inflammatory gene expression and inflammasome activation [11]. Collectively, excessive release of fragmented DNA, sourced from the nucleus (nDNA), or mitochondria (mtDNA), together with impaired clearance, may result in sustained cfDNA elevation, thereby amplifying innate immune responses and perpetuating inflammation through a positive feedback loop [12]. Interestingly, the nature of immune response to cfDNA seems to be sequence-dependent as suggested by the immunosuppressive effect of telomeric TTAGGG repetitive motif observed, for example, in human B cells [13].

In axSpA patients, serum levels of cfDNA were found to be higher in individuals with radiographic (r-) form of axSpA, even in those with axial involvement, compared to healthy subjects [14]. Likewise, levels of plasma cf-mtDNA were found to be elevated in r-axSpA patients with high disease activity determined by Ankylosing spondylitis disease activity score (ASDAS) with CRP > 2.1 [15]. When looking at mental health disorders, the body fluid DNA fragments, particularly plasma levels of cf-mtDNA, reflect psychosocial stress and depression [16] and its response to antidepressant treatment [17].

In axSpA, symptoms of depression and anxiety correspond to disease activity. Given the inflammatory background, we hypothesized a common association between axSpA disease activity and the severity of depression and anxiety. We looked for the plasma levels of DNA fragments – cf-nDNA and cf-mtDNA – in axSpA patients, and studied their relation to depression and anxiety. Further, we determined the abundance of telomeric repeats, and evaluated DNA integrity by calculating the DNA integrity index (DII).

## Methods

### Participants’ characteristics

Individuals with axSpA and healthy control counterparts (HCs), aged between 18 and 65 years, were included; all of them were participants of the RHEU-THYMIA study [18]. Written informed consent was required from all participants prior to all study procedures. The study was conducted in compliance with the Declaration of Helsinki and approved by the Ethical Committee of Rheumatology Institute in Prague (number 5704/2021).

The following data were gathered from all participants: sex, body mass index (BMI), comorbidities, pharmacotherapy, alcohol abuse, and physical activity in accordance with the World heath organization (WHO)’s weekly recommendations, Health assessment queationnaires (HAQ), and five-dimensional EuroQol questionnaire (EQ-5D) for quality of life [19]. The below exclusion criteria were applied: fibromyalgia (FM), other autoimmune disorders, chronic decompensated diseases e.g. type 2 diabetes, chronic lung, kidney and heart disease, malignancy, drug abuse, current infection, injuries or surgical care within the last 6 weeks, pregnancy, breastfeeding, and pharmacological therapy consisting of corticosteroids, opioid analgesics, any hormonal therapy including hormonal contraceptive or hormonal substitutive therapy, anti-seizure or anti-Parkinson’s disease drugs, monoclonal antibodies, cytokine analogues or recombinant proteins for any reason other than axSpA, or any experimental therapy.

All axSpA patients were required to fulfil the Assessment of Spondyloarthritis International Society (ASAS) classification criteria [20]. Based on SIJs radiographic findings, patients were classified as having r-axSpA or non-radiographic (nr)-axSpA. Categorical data (yes/no) regarding advanced disease, characterised by the presence of spinal involvement – specifically syndesmophytes – were collected. Characterisation of axSpA included current and previous treatments, disease duration, and the presence of inflammatory back pain (IBP), arthritis (in any of 44 joints), enthesitis (in any locations defined by the Maastricht AS enthesitis criteria), Bath AS metrology index (BASMI), and ASDAS-CRP [21]. Patient-reported outcomes (PROs) included the Bath Ankylosing Spondylitis Disease Activity Index (BASDAI) and the Bath AS Functional Index (BASFI).

### Mental health assessment

Mental disorders (MDs) were assessed using the Czech-validated version of the Mini-International Neuropsychiatric Interview (M.I.N.I.). Briefly, the M.I.N.I. is a structured diagnostic interview comprising modules corresponding to specific diagnostic categories and aligned with the Diagnostic and Statistical Manual of Mental Disorders, Fourth Edition (DSM-IV), and the International Classification of Diseases, Tenth Revision (ICD-10) criteria [22, 23]. Following initial screening questions, subsequent modules determine whether diagnostic criteria for each disorder are met. In the present analysis, positive modules for current or lifetime major depressive disorder, dysthymia, and suicidality were grouped as affective disorders (AffectD), while panic disorder and generalized anxiety disorder were classified as AnxD.

The validated Czech versions of the Beck Depression Inventory (BDI-II) and the Beck Anxiety Inventory (BAI) were utilized [24, 25]. Both inventories comprise 21 items that are rated on a four-point Likert scale (0–3). The BDI-II includes somatic, cognitive, and affective signs of depression, while the BAI covers anxiety symptoms such as dizziness, trembling, facial flushing, catastrophic thinking, and fear of death or loss of control. BDI-II scores indicate the following categories: no depression (0–13), mild (14–19), moderate (20–28), and severe (29–63) depression. BAI scores indicate the following categories: no anxiety (0–9), mild (10–18), moderate (19–29), and severe (30–63) anxiety [25, 26].

### Blood sampling and analyses

Blood samples were collected in the morning (08:00–12:30) following overnight fasting. The serum levels of CRP were analysed on the same day. All samples were immediately centrifuged, after which the plasma was stored at −70°C until analysis.

### Analysis of cf-nDNA, cf-mtDNA, T/S ratio, and DII

#### cfDNA Isolation from plasma

cfDNA was extracted from the plasma of axSpA patients and HCs using the QIAamp Circulating Nucleic Acid Kit (Qiagen) in accordance with the manufacturer’s protocol. The purified DNA was eluted in 150 μL of nuclease-free water, and its concentration and purity were determined by spectrophotometric measurement of absorbance at 260/280 nm using a NanoDrop ND-1000 UV/Vis spectrophotometer (Thermo Scientific). cfDNA samples were stored at 120°C until quantification, which was carried out within 24 hours of isolation.

#### cfDNA quantification

Standard curves were generated using serial dilutions of TaqMan Standard Human DNA (TaqMan Control Genomic DNA, Applied Biosystems, USA) for cf-nDNA and telomeric sequences; pure mitochondrial DNA extracted from whole blood of healthy donors not enrolled in the study was used to generate a standard curve for cf-mtDNA, following the protocol described by Ahmad et al. (2007) [27]. The DNA integrity index scored by ALU ratio analyzed short (ALU-115) and longer (ALU-247) ALU repetitive sequences; the standard curves for both ALU sequences were prepared using cfDNA isolated from the non-participants’ plasma pool. Triplicate qPCR reactions were performed, each in a total volume of 10 μL, comprising 5 μL qPCRBIO SyGreen Mix Separate-ROX (PCR Biosystems), 1 μL template DNA, nuclease-free water, and primers.

For nuclear cfDNA, a single-copy gene *RPLPO* encoding the acidic ribosomal phosphoprotein was amplified using a forward primer 5’-CAG CAA GTG GGA AGG TGT AAT CC-3’ and a reverse primer 5’-CCC ATT CTA TCA TCA ACG GGT ACA A-3’, both at 600 nM [28]. Mitochondrial cfDNA targeting the *MT-ND2* gene was amplified using a forward primer 5’-CACACTCATCACAGCGCTAA-3’ and a reverse primer 5’-GGATTATGGATGCGGTTGCT-3’ at 700 nM each [29]. For telomeric repeats, a forward primer 5’-ACACTAAGGTTTGGGTTTGGGTTTGGGTTTGGGTTAGTGT-3’, and a reverse primer 5’-TGTTAGGTATCCCTATCCCTATCCCTATCCCTATCCCTAACA-3’ [30], both at 500 nM, were employed. For ALU-115 amplification, a forward primer 5’-CCTGAGGTCAGGAGTTCGAG-3’ and a reverse primer 5’-CCCGAGTAGCTGGGATTACA-3’, both at 450 nM, and for ALU-247, a forward primer 51-GTGGCTCACGCCTGTAATC-31 and a reverse primer 51-CAGGCTGGAGTGCAGTGG-31 [31], both at 500 nM, were used.

Isolated cfDNA was quantified by using an absolute quantitative PCR (qPCR) performed on a LC480 LightCycler (Roche Applied, USA) using the following cycling program for cf-nDNA, cf-mtDNA, and both ALU sequences: initial denaturation at 951°C for 10 min, followed by 45 cycles of 951°C for 10 s (denaturation), 651°C for 10 s (annealing), and 721°C for 11 s (extension), concluding with a melting curve analysis from 60 to 971°C. For telomeric repeats, the cycle program started with a denaturation at 951°C for 2 min, followed by 50 cycles of 951°C for 5 s (annealing), and 551°C for 25 s (extension), concluding with a melting curve analysis from 50 to 951°C. Amplification efficiencies were 107% (slope −3.1708, R² 0.9998) for *RPLPO*, 105% (slope −3.2663, R² 0.9598) for *MT-ND2*, 96% (slope −3.4166, R² 0.9995) for ALU-115, 90% (slope −3.5993, R² 0.9991) for ALU-247, and 94% (slope −3.488, R² 0.9816) for telomeric repeats. Two concentrations from each standard curve series were included as inter-run calibrators on every PCR plate. To calculate the mean threshold cycle (Ct), an intra-assay error of 0.34 was applied: if the standard deviation from technical triplicate exceeded the intra-assay variation, outlier value was excluded from the further analysis. The T/S ratio reflects the amount of telomeric repeat sequences (T) in relation to nDNA established by quantifying a single-copy gene (S) [28]. The DII was expressed as the ALU-247/ALU-115 ratio.

### Statistical analysis

For descriptive statistical analysis continuous variables were presented as means with 95% CIs or medians with upper and lower quartiles while discrete variables as frequencies and percentages. Healthy controls were randomly frequency-matched to the patient group by age and sex. The t-test was used for univariate subgroup comparisons while the Wilcoxon exact or Mann-Whitney tests for non-parametric data. Correlations were performed using Spearman’s correlation coefficient. A p-value of less than 0.05 was considered statistically significant. Adjustment for multiple testing (Bonferroni correction) was applied where appropriate for predefined group comparisons. In contrast, biomarker and correlation analyses were considered exploratory; therefore, no adjustment for multiple testing was applied. Reported p-values are uncorrected and should be interpreted with caution as hypothesis-generating findings. To explore biomarker level contributions towards various outcomes (ASDAS-CRP level, BDI-II or BAI scores) we have employed linear regression modelling. Given the exploratory rather than predictive goal all models were constructed using all measured biomarkers. Mild heteroskedasticity was detected thus we have refitted them with weights computed as the inverse of the squared residuals as a commonly used heuristic (WLS). All biomarkers were log2 transformed prior modelling to account for non-linearity.

## Results

### Demographic data

Seventy-nine patients with axSpA and 88 age- and sex-matched HCs were included. Demographic and disease characteristics are presented in Table 1.

**Table 1.** Demographic data and axSpA characteristics.

| **Feature** | **axSpA (n=79)** | **HC (n=88)** |
|---|---|---|
| Age, median (IQR) | 41.00 (34.00 - 48.00) | 42.00 (28.75 - 48.50) |
| Sex – Female/Male, % | 35.44/64.56 | 29.55/70.45 |
| BMI, median (IQR) | 25.35 (23.11 - 29.08) | 26.66 (23.43 - 30.06) |
| Marital status – 1/2, % | 32.91/67.09 | 45.45/54.55 |
| Adults in household – =1/ >1, % | 22.78/77.22 | 26.14/73.86 |
| City population – <20k/20-100/>100k, % | 46.84/15.19/37.97 | 34.09/10.23/55.68 |
| Work status – 1/2/3, % | 86.10/10.10/3.80 | 80.70/4.60/14.80 |
| HAQ, median (IQR) | 0.12 (0.00 - 0.62)* | 0.00 (0.00 - 0.00)* |
| 5D-EQ, median (IQR) | 0.80 (0.69 - 0.80)* | 1.00 (0.83 - 1.00)* |
| Comorbidities – untracked dis./up to 2/>2, % | 54.43/31.65/5.06 | 57.95/37.50/4.55 |
| Alcohol consumption below/in line/above WHO recommendations<sup>a</sup>, % | 84.81/12.66/2.53 | 80.68/13.64/5.68 |
| Meets WHO physical activity recommendations<sup>b</sup>, % | 68.35 | 64.77 |
| Disease duration since first symtom, median (IQR) | 12.22 (6.36 - 18.42) |  |
| Disease duration since diagnosis, median (IQR) | 6.16 (2.91 - 10.07) |  |
| HLA B27 positive, % | 92.41 |  |
| Non-radiographic axSpA, % | 39.24 |  |
| Radiographic axSpA, % | 60.76 |  |
| SIJ involvement only, % | 67.10 |  |
| Spine involvement, % | 32.90 |  |
| Hip involvement, % | 26.58 |  |
| Current peripheral arthritis/enthesitis/dactylitis, % | 11.39/17.72/3.80 |  |
| CRP, median (IQR) | 2.11 (1.17 - 6.90) |  |
| ASDAS-CRP, median (IQR) | 1.75 (1.15 - 2.76) |  |
| ASDAS-CRP, without activity (0-1.3), % | 30.38 |  |
| ASDAS-CRP, mild activity (1.4-2.0), % | 32.91 |  |
| ASDAS-CRP, moderate activity (2.1-3.4), % | 25.32 |  |
| ASDAS-CRP, severe activity (>3.5), % | 11.39 |  |
| BASDAI, median (IQR) | 2.80 (1.40 - 4.30) |  |
| BASFI, median (IQR) | 1.30 (0.20 - 3.00) |  |
| Medication – bDMARDs, % | 18.99 |  |
| Medication – csDMARDs, % | 8.86 |  |

Abbreviations: axSpA – Axial spondyloarthritis; HC – Healthy controls; BMI – Body mass index; Marital status: 1 – married/partner, 2 – single, divorced, widowed; WHO – World health organization; HAQ – Health assessment questionnaire; 5D-EQ – Five dimensional quality of life; Work status: 1 – full-time/part-time; 2 – sick-leave/unemployed/diseased pension; 3 – student/parental-leave/pension; IBP – Inflammatory back pain; CRP – C reactive protein; ASDAS-CRP – Ankylosing spondylitis (AS) disease activity score with CRP; BASDAI **–** Bath AS disease activity index; BASFI **–** Bath AS functional index; BASMI – Bath AS mobility index; SIJ – Sacroiliac joint; DMARDs – Disease modifying drugs; b – Biologic; cs – Conventional synthetic.
<sup>a</sup> WHO Recommendation for alcohol consumption: 2 standard units for women and 3 standard units for men; standard units: hard alcohol – 0.04l shot 1.3 units, beer 10° 0.33l 1 unit, beer 12° 1.5 units, wine 1dcl 1 unit.
<sup>b</sup> WHO Physical activity: 30 minutes of moderate-intensity physical activity 5 times a week or 20 minutes of high-intensity physical activity 3 times a week.
Statistical significance: \*p < 0.0001

Within the axSpA cohort, significant sex differences were observed in following disease-related variables: the distribution of non-radiographic versus radiographic axSpA (60.71%/39.29% in females vs. 27.45%/72.55% in males; p < 0.01), disease limited to the SIJ versus spinal involvement (89.30%/10.70% vs. 54.90%/45.10%, respectively; p < 0.001), and hip involvement (10.71% vs. 35.29%, respectively; p < 0.01) (Supplementary Table S1). In addition, males more frequently met WHO recommendations for physical activity and were more often treated with bDMARDs than females (80.39% vs. 46.43%, respectively; p < 0.01, and 27.45% vs. 3.57%, respectively; p < 0.01) (Supplementary Table S1).

### Mental burden in axSpA

Patients with axSpA experienced affective and anxiety disorders more frequently than HC (26.58% vs. 2.27%, p < 0.0001 and 13.92% vs. 4.55%, p < 0.01, respectively). No significant sex differences were observed within the axSpA cohort; however, both female and male patients with axSpA differed significantly from their healthy counterparts. Depressive symptoms assessed by the BDI-II were present in 20.25% of patients with axSpA compared with 2.27% of HCs (p < 0.01), with similar prevalence across sexes. Somatic and affective dimensions contributed most prominently to depressive symptomatology, particularly in male patients with axSpA (Supplementary Table S2). With respect to disease phenotype, disease activity rather than radiographic pattern was associated with mental health burden. Mental disorders, depressive symptoms, and anxiety – assessed using the M.I.N.I., BDI-II, and BAI – were significantly more frequent in patients with active disease compared with those with inactive axSpA (MD according to M.I.N.I.: 44.00% vs. 13.79%, p < 0.01; BDI-II: 30.00% vs. 3.45%, p < 0.0001; BAI: 34.00% vs. 3.45%, p < 0.001, respectively). Differences were mainly driven by somatic and affective components of the BDI-II (Supplementary Table S3).

### Variability of cf-DNA in axSpA and across sexes

The plasma levels of different types of DNA fragments in axSpA and HC are demonstrated in Figure 1. Among the assessed cfDNA parameters, patients with axSpA exhibited significantly higher plasma levels of cf-mtDNA compared to HCs [2.23ng/ml (1.14 - 4.36) vs. 1.68ng/ml (1.02 - 2.55), p < 0.05]. In contrast, DNA integrity was significantly lower in axSpA than in HCs [1.03 (0.84 - 1.39) vs. 1.26 (0.96 - 1.42), p < 0.05]. The levels of cf-nDNA tended to be lower in patients with axSpA compared with HCs, although this difference did not reach statistical significance (p = 0.07).

**Figure 1.**
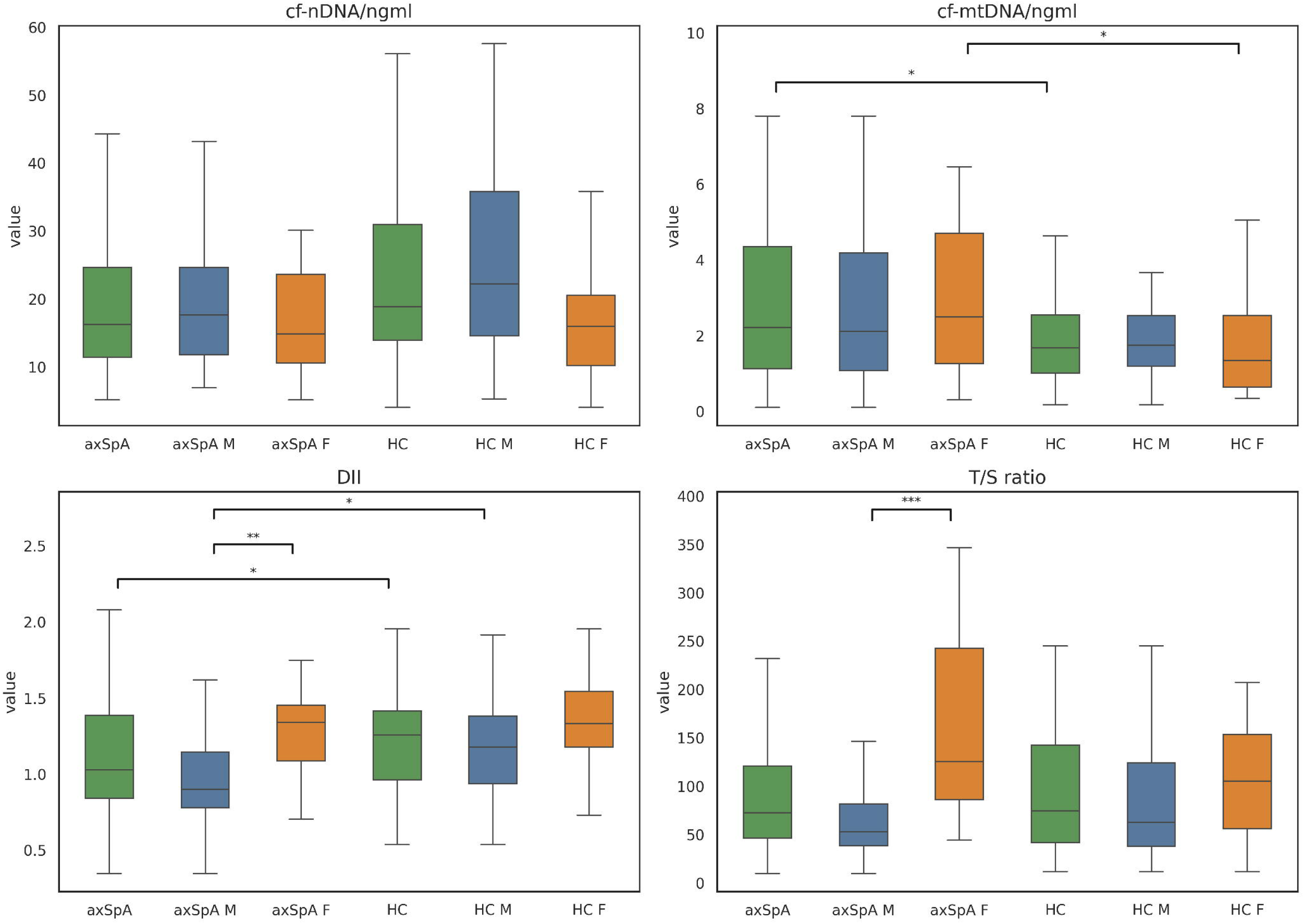
Variability in plasma levels of cf-nDNA, cf-mtDNA, DII, and T/S ratio in patients with axSpA and healthy controls. Abbreviations: cf-n/mtDNA – Cell-free circulating nuclear/mitochondrial DNA; DII – DNA integrity index; T/S – Telomere-to-single copy gene; axSpA – axial spondyloarthriti;, HC – healthy controls; M-males; F – females. Statistical significance: *p < 0.05; **p < 0.01; ***p < 0.001; P-values are uncorrected; no adjustment for multiple testing was applied due to the exploratory nature of the analyses.

As illustrated in Figure 1 and Supplementary Table S4, pronounced sex-related differences were observed. The DII was significantly lower in males with axSpA compared with females with axSpA and healthy males, whereas cf-mtDNA levels were significantly higher in females with axSpA compared with their respective counterparts.

### Relationships between cf-nDNA and cf-mtDNA

Although cf-nDNA levels positively correlated with cf-mtDNA in both groups, patients with axSpA and HC, (Table 2; Supplementary Table S5) there were sex dependent differences. These associations were particularly pronounced in the HC group, with a strong positive correlation between cf-nDNA and cf-mtDNA observed in females (r_s_ = 0.618, p < 0.001; Supplementary Table S6). In contrast, in patients with axSpA, a significant positive correlation between cf-nDNA and cf-mtDNA (r_s_ = 0.441, p < 0.05) was observed in males and weaker in females (r_s_ = 0.398, p < 0.05) (Supplementary Table S7).

**Table 2.** Spearman’s correlation of DNA fragments and clinical variables in axSpA patients.

|  | cf-nDNA | cf-mtDNA | DII | T/S ratio |
|---|---|---|---|---|
| cf-nDNA | 1 | ***0,430 | -0,114 | ***-0,563 |
| cf-mtDNA | ***0,430 | 1 | -0,192 | -0,147 |
| DII | -0,113 | -0,192 | 1 | ***0,462 |
| T/S ratio | ***-0,563 | -0,147 | ***0,462 | 1 |
| BMI | *0,282 | 0,043 | -0,086 | *-0,259 |
| Comorbidities | 0,099 | 0,049 | 0,053 | -0,173 |
| WHO Physical activity | -0,007 | -0,136 | -0,198 | *-0,267 |
| WHO Alcohol consumption | -0,010 | 0,024 | 0,028 | -0,115 |
| HAQ | 0,189 | 0,058 | -0,090 | -0,189 |
| 5D-EQ | -0,050 | 0,075 | -0,038 | 0,140 |
| MD (M.I.N.I.) | -0,197 | -0,115 | 0,008 | 0,113 |
| BDI-II | 0,041 | -0,026 | -0,055 | -0,131 |
| BAI | 0,035 | -0,111 | 0,133 | 0,014 |
| BDI-II Somatic scores | 0,029 | 0,023 | 0,025 | -0,107 |
| BDI-II Cognitive scores | -0,090 | -0,045 | -0,081 | -0,024 |
| BDI-II Affective scores | 0,066 | -0,165 | -0,055 | -0,141 |
| DisDur since symptoms | 0,056 | -0,121 | -0,102 | -0,172 |
| DisDur since diagnosis | 0,103 | -0,021 | 0,039 | -0,044 |
| Radiographic axSpA | 0,113 | -0,188 | -0,210 | *-0,257 |
| Spine involvement | 0,147 | -0,125 | *-0,231 | *-0,279 |
| CRP | 0,180 | -0,017 | *-0,244 | **-0,371 |
| ASDAS-CRP | 0,011 | -0,066 | -0,044 | -0,190 |
| BASDAI | -0,045 | -0,024 | 0,105 | -0,053 |
| BASFI | 0,158 | 0,062 | -0,095 | **-0,295 |
| Medication bDMARDs | -0,187 | -0,058 | 0,174 | 0,155 |
| HLA-B27 positivity | 0,133 | 0,061 | -0,006 | -0,080 |
| Male sex | 0,110 | -0,053 | **-0,381 | ***-0,520 |
| Age | 0,160 | 0,082 | -0,063 | *-0,223 |

Abbreviations: axSpA – Axial spondyloarthritis; cf-n/mtDNA – Cell-free circulating nuclear/mitochondrial DNA; DII – DNA integrity index; T/S – Telomere-to-single copy gene; BMI – Body mass index; WHO – World health organization; HAQ – Health assessment questionnaire; 5D-EQ – Five dimensional quality of life; MD – Mental disorder according to M.I.N.I. (Mini-International Neuropsychiatric Interview); BDI-II – Beck depression inventory-II; BAI – Beck anxiety inventory; DisDur – Disease duration; CRP – C reactive protein; ASDAS-CRP – Ankylosing spondylitis (AS) disease activity score with CRP; BASDAI – Bath AS disease activity index; BASFI – Bath AS functional index; DMARDs – Disease modifying drugs; b – Biologic; HLA – Human leukocyte antigen.
Statistical significance: \* p<0.05, \*\* p<0.01, \*\*\* p<0.0001
Data are presented as Spearman’s correlation coefficients (rho). Statistical significance is indicated as follows: \* p < 0.05; \*\* p < 0.01; \*\*\* p < 0.0001. Due to the exploratory nature of the biomarker analyses, no adjustment for multiple testing was applied. Reported p-values are uncorrected and should be interpreted with caution as hypothesis-generating findings.

### Mental health burden and circulating DNA fragment levels

In axSpA, no significant associations were observed between the presence of mental disorders, depression, or anxiety and levels of cf-nDNA, cf-mtDNA, T/S ratio, or DNA integrity (Table 3; Supplementary Tables S5, S7). In healthy males, however, BDI-II and its somatic subscale scores showed a negative correlation with T/S ratio (r_s_ = −0.280 and r_s_ = −0.316, both p < 0.05, see Supplementary Table S6).

**Table 3.** Plasma levels in the circulating DNA fragments in axSpA (n=79)

|  | cf-nDNA (ng/ml) | cf-mtDNA(ng/ml) | DII | T/S ratio |
|---|---|---|---|---|
| r-axSpA (n=48) | 18.49 (11.06 - 25.25) | 1.99 (1.02 - 4.05) | 0.95 (0.78 - 1.33) | 62.50 (45.41 - 103.69) * |
| nr-axSpA (n=31) | 14.37 (12.17 - 20.92) | 2.89 (1.43 - 5.74) | 1.14 (0.91 - 1.41) | 96.25 (54.66-219.22) |
| SIJ limited (n=53) | 14.37 (10.70 - 25.14) | 2.26 (1.27 - 4.84) | 1.14 (0.89 - 1.40) | 82.77 (51.13 - 148.96) † |
| Spine involvement (n=26) | 20.32 (12.77 - 24.42) | 2.05 (1.08 - 3.49) | 0.88 (0.77 - 1.14) | 53.48 (34.33 - 85.25) |
| Active disease (ASDAS-CRP ≥ 1.4) (n=50) | 16.98 (10.66 - 24.42) | 2.06 (1.08 - 4.18) | 1.06 (0.83 - 1.39) | 66.14 (46.56 - 100.43) |
| Inactive disease (ASDAS-CRP < 1.4) (n=29) | 15.77 (11.86 - 25.80) | 2.42 (1.37 - 4.62) | 1.02 (0.87 - 1.35) | 101.90 (51.13 - 146.63) |
| Overweight (BMI ≥ 25) (n=42) | 19.88 (12.75 - 26.18) | 2.19 (1.36 - 3.50) | 0.97 (0.82 - 1.34) | 66.14 (38.69 - 110.83) |
| Normal weight (BMI < 25) (n=37) | 14.37 (10.31 - 19.31) | 2.23 (1.03 - 5.40) | 1.12 (0.89 - 1.41) | 81.50 (51.13 - 156.36) |
| Depression (BDI-II) (n=16) | 12.17 (9.54 - 21.40) | 2.12 (0.91 - 3.29) | 1.14 (0.82 - 1.40) | 71.08 (41.49 - 115.47) |
| Without Depression (BDI-II) (n=63) | 17.03 (12.22 - 24.99) | 2.26 (1.21 - 4.43) | 1.02 (0.85 - 1.36) | 72.95 (47.54 - 121.02) |
| Anxiety (BAI) (n=18) | 13.97 (9.55 - 25.02) | 1.89 (0.73 - 3.31) | 1.25 (0.90 - 1.41) | 94.33 (48.54 - 143.69) |
| Without Anxiety (BAI) (n=61) | 17.03 (11.94 - 23.75) | 2.26 (1.25 - 4.62) | 1.01 (0.83 - 1.33) | 72.44 (46.40 - 111.12) |
| M.I.N.I. AffectD (n=21) | 14.06 (9.57 - 23.58) | 2.02 (1.00 - 4.49) | 1.32 (0.86 - 1.41) | 89.02 (49.73 - 148.96) |
| M.I.N.I. Without AffectD (n=58) | 16.98 (11.96 - 25.56) | 2.26 (1.19 - 4.11) | 1.01 (0.84 - 1.32) | 68.66 (46.56 - 110.83) |
| M.I.N.I. AnxD (n=11) | 15.88 (12.84 - 24.11) | 1.89 (0.97 - 4.66) | 1.36 (0.81 - 1.46) | 67.83 (51.69 - 113.77) |
| M.I.N.I. Without AnxD (n=68) | 16.35 (11.12 - 25.07) | 2.24 (1.15 - 4.16) | 1.02 (0.86 - 1.35) | 73.45 (46.34 - 119.76) |

Abbreviations: cf-n/mtDNA – Cell-free circulating nuclear/mitochondrial DNA; DII – DNA integrity index; T/S – Telomere-to-single copy gene; r/nr-axSpA – Radiographic/non-radiographic axial spondyloarthritis; SIJ – Sacroiliac joints; ASDAS-CRP – Ankylosing spondylitis disease activity score with C-reactive protein; BMI – Body mass index; BDI-II – Beck depression inventory; BAI – Beck anxiety inventory; M.I.N.I. – Mini-International Neuropsychiatric Interview; AffectD – Affective disorders; AnxD – Anxiety disorders.
Different symbols indicate statistically significant differences (p < 0.05) for specific comparisons:
\* r-axSpA vs nr-axSpA; † axSpA with sacroiliac joint involvement only vs spinal involvement.
P-values are uncorrected; no adjustment for multiple testing was applied due to the exploratory nature of the analyses.

The associations of cf-nDNA, cf-mtDNA, T/S ratio and DII with depressive and anxiety symptoms are shown in Figure 2A and 2B and Supplementary Table S8. In patients with axSpA, higher DNA integrity index was significantly associated with greater anxiety severity, with a one-unit increase in DII corresponding to a 1.40-point increase in BAI score (*coeff. 1.397, SE 0.447, p < 0.01*). A similar trend was observed for depressive symptoms, although the association did not reach statistical significance (*coeff. 1.032, SE 0.535, p = 0.06*). CRP showed the strongest association with both psychological outcomes, as a doubling of CRP levels was associated with an increase of 4.26 points in BDI-II and 4.01 points in BAI scores. In contrast, both cf-nDNA and cf-mtDNA showed only minor associations with anxiety severity (*coeff. −0.032, SE 0.016, p < 0.05 and coeff. −0.246, SE 0.070, p < 0.001,* respectively). Additionally, cf-nDNA levels tended to be lower in patients with axSpA and AffectD, although this finding did not reach statistical significance (p = 0.07, Table 3).

**Figure 2.**
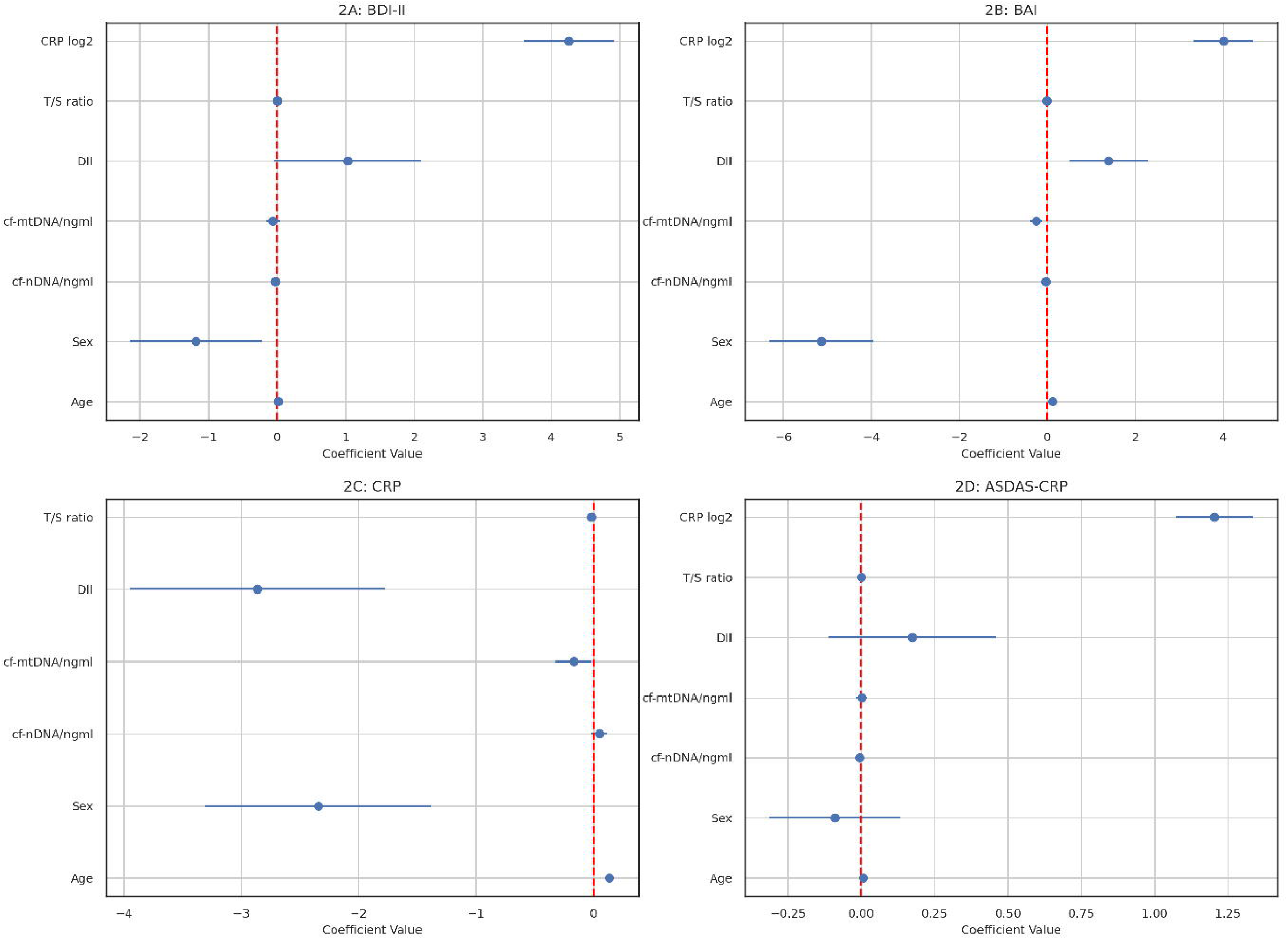
Linear regression models for mental health, disease activity and cf-DNA in axSpA. Abbreviations: axSpA – Axial spondyloarthritis; BAI – Beck anxiety inventory; BDI-II – Beck depression inventory-II; CRP – C reactive protein; ASDAS-CRP – Ankylosing spondylitis (AS) disease activity score with CRP; cf-n/mtDNA – Cell-free circulating nuclear/mitochondrial DNA; DII – DNA integrity index; T/S – Telomere-to-single copy gene.

### Circulating DNA fragment levels in relation to clinical features of axSpA

Among the assessed clinical parameters, CRP levels and radiographic phenotype were associated with circulating plasma DNA fragment parameters. Serum CRP levels negatively correlated with both DII and relative T/S ratio (Table 2), while the association between T/S ratio and CRP was observed in both sexes (Supplementary Table S7). In contrast, none of the circulating DNA parameters differed between patients with active and inactive disease (Table 3), and no significant associations with ASDAS-CRP or BASDAI were identified (Table 2).

We further evaluated the effects of cf-nDNA, cf-mtDNA, T/S ratio and DII on CRP, disease activity (Figure 2C, 2D and Supplementary materiál Table S8) and radiographic phenotype. Linear regression modeling demonstrated that DII significantly negatively influences the CRP levels: a one-unit increase in DII is associated with a decrease of 2.86 units in CRP (*coeff. −2.861, SE 0.544, p < 0.001).* The significant negative effect of cf-mtDNA and T/S ratio was also observable, but the effect size was limited (*coeff. −0.165 SE 0.077*, *p < 0.05 and coeff. −0.016 SE 0.002 p < 0.001, respectively*), see Figure 2C. Index ASDAS-CRP was significantly associated with CRP levels (*coeff. 1.205, SE 0.066, p < 0.001)* and only marginaly with cf-nDNA (*coeff. −0.060, SE 0.003, p < 0.05),* see Figure 2C.

With respect to radiographic phenotype, patients with radiographic axSpA and spinal involvement exhibited significantly lower T/S ratio levels compared with patients without radiographic changes (Table 3). Consistently, T/S ratio weakly negatively correlated with both r-axSpA phenotype and spinal involvement (Table 2). The multiple regression model to develop r-axSpA rather than nr-axSpA is demontrated in Table 4. The cf-mtDNA emerged as a negative predictor (*coeff −0.231 OR 0.79, p < 0.05*), whereas male sex showed a trend toward increased risk (*coeff. 1.176 OR 3.24, p = 0.06*).

**Table 4.** Multiple regression model for presence of r-axSpA.

|  | **coef.** | **OR** | **p** | **5%** | **95%** |
|---|---|---|---|---|---|
| Age | 0.008 | 1.01 | 0.795 | -0.044 | 0.060 |
| Male sex | 1.176 | 3.24 | 0.063 | -0.068 | 2.419 |
| cf-nDNA | 0.042 | 1.04 | 0.187 | -0.020 | 0.103 |
| cf-mtDNA | -0.232 | 0.79 | 0.020 | -0.427 | -0.036 |
| DII | -0.505 | 0.60 | 0.514 | -2.022 | 1.012 |
| T/S ratio | -0.001 | 1.00 | 0.621 | -0.008 | 0.005 |
| CRP | 0.489 | 1.63 | 0.232 | -0.313 | 1.292 |
| Intercept | -0.669 | 0.51 | 0.734 | -4.536 | 3.196 |

Abbreviations: r-axSpA – Radiographic axial spondyloarthritis; cf-n/mtDNA – Cell-free circulating nuclear/mitochondrial DNA; DII – DNA integrity index; T/S – Telomere-to-single copy gene; CRP – C reactive protein.

We then examined whether cfDNA was associated with the other clinical parameters. Correlation analyses further revealed a negative association between physical activity and relative abundance of telomeric repeats in axSpA patients (Table 2), whereas no such relationship was observed in HC (Supplementary Table S5). BMI positively correlated with DII in HC, particularly males (r_s_ = 0.299 and 0.361, respectively, p < 0.01 and comorbidities significantly negatively correlated with T/S ratio in both sexes with r_s_ varying between −0.237 to −0.420 (p < 0.05), see Supplemmentary Table S5 and S6, but similar findings were not find in axSpA patients.

## Discussion

To our knowledge, this study provides the first comprehensive insight into the relationships between circulating DNA fragments in patients with axSpA, with particular attention to mental health burden. Although the findings should be interpreted in light of several limitations, our data suggest that circulating cell-free DNA exerts only a marginal effect on mood and anxiety symptoms in axSpA compared with the dominant influence of CRP. Nevertheless, cf-nDNA and cf-mtDNA were closely interrelated, and relative abundance of telomeric repeats and the level of DNA integrity varied by sex in axSpA and may influence disease-related outcomes.

In our cohort, the burden of depression and anxiety was higher in patients with axSpA and was substantially associated with disease activity, but not with radiographic pattern. Psychological burden affected both sexes; however, male sex appeared to be protective against depression and anxiety, in line with data from the general population [32]. Notably, males with axSpA in our cohort were considerably more affected than their healthy counterparts, particularly with respect to the severity of depressive symptoms. This finding warrants attention in clinical practice, given the impact of axSpA on psychosocial functioning in men and the higher prevalence of axSpA among middle-aged males.

Our hypothesis regarding a potential role of circulating DNA fragments in depression and anxiety in axSpA was based on the concept of neuroinflammation [9, 10, 33], particularly the immunostimulatory properties of cf-mtDNA [12]. In the present study, however, only DNA integrity level appeared to be associated with anxiety severity, whereas cf-mtDNA and cf-nDNA showed only marginal effects. Although altered plasma levels of cf-mtDNA have been reported in individuals with MDD [16, 17], other studies failed to confirm this association [34, 35]. Moreover, a recent meta-analysis including patients with mental and somatic disorders suggested that elevated plasma cf-mtDNA reflects psychological stress primarily in individuals with MDD and bipolar affective disorder, but not in those with somatic diseases such as diabetes [36]. Potential mechanisms linking cf-mtDNA to depression may involve its immunomodulatory properties and structural integrity. In patients with MDD, mtDNA copy number has been shown to correlate with interleukin (IL)-4, but not with IL-6 [37]. At the same time, MDD has been consistently associated with elevated levels of pro-inflammatory cytokines, including IL-6, IL-12, IL-18, tumour necrosis factor (TNF)-α, and CRP, accompanied by reduced levels of the anti-inflammatory cytokine IL-4 [38]. In patients with late-life depression, a higher deletion rate of cf-mtDNA has been linked to greater depressive symptom severity and was associated with IL-6 levels [39]. As we confirm the close link between inflammation (CRP levels), disease activity of axSpA, and mental burden, identifying immune mechanisms to develop depression or anxiety in axSpA patients is warranted.

In axSpA, cf-mtDNA levels were higher in our study; however, no association with disease activity was observed. Although cf-nDNA and cf-mtDNA were positively correlated, their association with CRP or ASDAS-CRP, which would be clinically relevant, was rather low or none. In contrast, a previous study by Peng et al. reported elevated cfDNA levels in patients with shorter disease duration, higher levels in males, and positive correlations with CRP and ASDAS-CRP [14]. Differences between serum and plasma cf-mtDNA measurements may partly explain these discrepancies, as serum cf-mtDNA levels are likely influenced by platelet activation [40], and platelet counts are known to increase during active disease [41]. Compared to our findings for cf-nDNA and cf-mtDNA, both correlation analysis and a regression model revealed a negative association between CRP and DII, and CRP and T/S ratio. Calculated as the ratio of ALU-247 bp to ALU-115 bp fragments, DII is a widely used probabilistic indicator of a relative increase in longer cfDNA fragments, which likely reflects a greater contribution from necrotic or other non-apoptotic cell death pathways rather than apoptosis alone [42]. We found that an increased inflammatory response, as indicated by higher levels of CRP, is associated with a decreased DII, which may indicate a shift toward a more frequent occurence of apoptotic cell death. This aligns with prior evidence that CRP can induce apoptosis in vascular smooth muscle cells [43]. In this context it is also noteworthy that CRP has been shown to facilitate the noninflammatory clearance of apoptotic cells [44]. Higher CRP associated with lower T/S ratio in axSpA patients corresponds with a recent finding of Liu et al. (2025) who showed that elevated CRP relative to circulating lymphocyte counts were consistently associated with shorter leukocyte telomeres [45].

In our cohort, associations were observed predominantly with more advanced disease features. Although no relationship with disease duration was detected, patients with spinal involvement and radiographic sacroiliitis exhibited lower T/S ratio. This would implicate lower abundance of telomeric sequences relative to single-copy gene, and eventually suggest weaker immunoinhibition mediated by circulating cfDNA in comparison to nr-axSpA subjects. Given the specific clinical and immunological features of axSpA, it is worth noting that telomeric sequences present in native plasma and serum were shown – at least in vitro – to inhibit TNF-α mRNA expression in monocytes [28]. Moreover, the T/S ratio is proportional to the average telomere length (TL), which shortens with age or environmental and lifestyle-related factors, including BMI or stress [46], and the presence of chronic inflammatory disease and cumulative inflammatory burden [47]. All of this could help explain the negative association of T/S with more pronounced disease characteristics.

In a regression model, cf-mtDNA emerged as a clinically rather negative predictor of non-radiographic / radiographic feature of axSpA; in contrast, DII appears to be a clinically more promising (albeit statistically non-significant) predictor of a radiographic involvement, which could suggest that the lower DII, the higher probability of radigrafic manifestation of axSpA. This finding goes well with our other observation of lower DII in men than in women and, finally, higher risk of developing r-axSpA in males than females. However, the present study was primarily focused on mental health outcomes and disease activity, and therefore, only categorical data on sacroiliac joint and spinal involvement were available, without quantitative scores for inflammation or structural damage. While it may be speculated that increased cell death and subsequent release of cf-mtDNA occur in the early phases of axSpA, further dedicated studies are required to address this hypothesis. Recent data have also implicated activation of the innate immune system, including components of the complement cascade, in radiographic progression [48].

Finally, we observed substantial sex-related differences in T/S ratio and DNA integrity in axSpA. Telomere length was found to be associated with estrogen [49], thus accounting for sex specific shortening. Next to that, in axSpA, sex-specific differences in CRP levels and mobility have previously been reported [50], and female patients may experience longer diagnostic delay due to the absence of radiographic sacroiliitis, whereas male sex has been associated with greater spinal radiographic progression [1]. Thus, all these factors may contribute to the observed sex-specific patterns in circulating DNA fragment profiles.

## Limits of the study

Several limitations should be considered when interpreting the results of this study. First, the cross-sectional design precludes assessment of temporal relationships and prevents evaluation of changes in mental health or disease activity over time or in response to therapeutic interventions. Second, given the rapid degradation of circulating free DNA, the reliance on a single time-point blood sampling represents an additional limitation. Although sample collection followed clinical assessment, pre-analytical conditions prior to blood draw could not be fully standardised.

A further limitation is the availability of only categorical data regarding radiographic disease characteristics. Consequently, findings related to radiographic patterns should be interpreted as exploratory and will require confirmation in future studies, preferably with longitudinal design and detailed quantitative imaging assessments.

## Conclusion

In conclusion, this cross-sectional observational analysis identified sex-related differences in several disease-associated parameters, including the distribution of non-radiographic vs. radiographic axSpA, SIJ vs. spinal involvement, or hip involvement. Sex differences were also observed in the patient cohort in relation to the burden of depression and anxiety. Furthermore, patients with axSpA exhibited a greater mental burden than healthy individuals, which was associated with disease activity.

The examination of the role of circulating cf-nDNA, cf-mtDNA, abundance of telomeric repeats and integrity of DNA in mental health revealed only a marginal effect on mood and anxiety symptoms in axSpA compared with the dominant influence of CRP. No association was found between cf-mtDNA and depression or anxiety; instead, DNA integrity was associated with anxiety severity. In contrast to mental burden, axSpA clinical parameters including CRP levels and radiographic pattern were associated with plasma cfDNA, particularly the relative abundance of telomeric sequences and DNA integrity. Finally, cfDNA differences by sex and axSpA disease status were observed. These results suggest that circulating fragments of cfDNA may play a role primarily in the early stages of axSpA and warrant further investigation into their role in axSpA manifestation and progression, and potential as biomarkers.

## Data Availability

The datasets generated during the current study are not publicly available due to clinical data but are available from the corresponding author on reasonable request.

## List of abbreviations

axSpA: Axial spondyloarthritis
cf-nDNA: Cell-free nuclear DNA
cf-mtDNA: Mitochondrial DNA
T/S ratio: Relative telomere length
DII: DNA integrity index
HC: Healthy controls
M.I.N.I.: Mini-International Neuropsychiatric Interview
BDI-II: Beck Depression Inventory-II
BAI: Beck Anxiety Inventory
BASDAI: Bath Ankylosing Spondylitis Disease Activity Index
CRP: C-reactive protein
ASDAS-CRP: Ankylosing Spondylitis Disease Activity Score with C-reactive protein
SIJs: Sacroiliac joints
MDD: Major depression RR- Relative risk
DI: Confidence interval
AnxD: Anxiety disorders
HR: Hazard ratio
IMAS: The International Map of Axial Spondyloarthritis
BMI: Body mass index
WHO: World heath organization
HAQ: Health assessment queationnaires
EQ-5D: Five-dimensional EuroQol questionnaire
ASAS: Assessment of Spondyloarthritis International Society
IBP: Inflammatory back pain
MASES: Maastricht AS enthesitis criteria
BASMI: Bath AS metrology index
PROs: Patient-reported outcomes
BASFI: Bath AS Functional Index
DSM-IV: Diagnostic and Statistical Manual of Mental Disorders, Fourth Edition
ICD-10: International Classification of Diseases, Tenth Revision
IL: Interleukin
TNF-α: Tumour necrosis factor-α

## Acknowledgements

Authors would like to thank Mrs. Alena Swienteková, Mrs. Barbora Smetanová, Mrs. Eliška Vejvodová, Mrs. Jiřina Kvítková, Mrs. Tetyana Napuda and Mr. Martin Bednář for technical support.

## Group Authorship List

M.H., K.P., K.K., P.D. - Conceptualization, P.M., M. O., M.H., K.P., K.K, P.D. - Methodology, M.O., M.H., K.P. - Patients investigation, B.B., P.K. P.D. - laboratory analysis, M.H., J.K., L.S. - Formal analysis and investigation, M.H., B.B.- Writing - original draft preparation,P.M., M. O., M.H., K.P., K.K, P.D., P.S, K.K., J.K.- Writing - review and editing; M.H., K.P. - Funding acquisition

## Disclouser

### Ethical Approval

All participants agreed to take part in the study and followed the Informed Consent approved by the Ethics Committee of the Institute of Rheumatology in Prague (No. 5704/2021).

### Consent for publication

Not applicable.

### Conflict of Interest Statement

The authors declare that they have no conflicts of interest regarding the present manuscript. All support received in the previous three years:

Research grant dedicated to the Institute of Rheumatology in Prague: **MH** - NU21-09-0097; **KP** - NU23-10-00434

Research grants Czech Health Research Council (NIMH), the Czech Research Foundation (NIMH), and the European Brain Council: **PM**

Meeting supports: **MH** – UCB

Honoraria for lecture and speakers fees: **KP** - AbbVie, Pfizer, Eli Lilly, MSD, Biogen, UCB, **LŠ** - AbbVie, AstraZeneca, Bristol-Myers Squibb, Eli Lilly, GlaxoSmithKline, Johnson & Johnson, Merck Sharp and Dohme, Novartis, Sanofi, Sandoz, Swedish Orphan Biovitrum, Swixx Biopharma, UCB; **PM** - Johnson & Johnson, Lundbeck, Richter Gedeon, AstraZeneca, Angelini, Teva, Boehringer Ingelheim, KRka

All other authors also declare that they have no conflicts of interest.

## References

1. Navarro-Compán, V., et al., Axial spondyloarthritis. Lancet, 2025. 405(10473): p. 159–172.

2. Rosine, N., et al., T cells in the pathogenesis of axial spondyloarthritis. Joint Bone Spine, 2023. 90(6): p. 105619.

3. Poddubnyy, D., et al., Psychosocial burden of axial spondyloarthritis and impact of different disease domains: a systematic literature review. Rheumatol Adv Pract, 2025. 9(3): p. rkaf063.

4. Park, J.Y., et al., The incidence of depression and anxiety in patients with ankylosing spondylitis: a systematic review and meta-analysis. BMC Rheumatol, 2020. 4: p. 12.

5. Shen, C.C., et al., Risk of Psychiatric Disorders following Ankylosing Spondylitis: A Nationwide Population-based Retrospective Cohort Study. J Rheumatol, 2016. 43(3): p. 625–31.

6. Garrido-Cumbrera, M., et al., Factors Associated with Poor Mental Health in Patients with Axial Spondyloarthritis: Results from the International Map of Axial Spondyloarthritis (IMAS). RMD Open, 2024. 10(2).

7. Zhao, S., et al., The prevalence of depression in axial spondyloarthritis and its association with disease activity: a systematic review and meta-analysis. Arthritis Res Ther, 2018. 20(1): p. 140.

8. Zhao, S.S., et al., Disease activity and mental health symptoms in axial spondyloarthritis: concordant or discordant? Rheumatology (Oxford), 2025.

9. Frank, P., et al., Association Between Systemic Inflammation and Individual Symptoms of Depression: A Pooled Analysis of 15 Population-Based Cohort Studies. Am J Psychiatry, 2021. 178(12): p. 1107–1118.

10. Wessa, C., M. Simon, and L. De Picker, Current evidence on immune-driven depression. Curr Opin Psychiatry, 2025.

11. Liu, F., et al., Cell-free DNA: a metabolic byproduct with diagnostic and prognostic potential in rheumatic disorders. Front Pharmacol, 2025. 16: p. 1537934.

12. Kustanovich, A., et al., Life and death of circulating cell-free DNA. Cancer Biol Ther, 2019. 20(8): p. 1057–1067.

13. Duramad, O., et al., Inhibitors of TLR-9 act on multiple cell subsets in mouse and man in vitro and prevent death in vivo from systemic inflammation. J Immunol, 2005. 174(9): p. 5193–200.

14. Peng, Y., et al., Circulating cell-free DNA correlate to disease activity and treatment response of patients with radiographic axial spondyloarthritis. Sci Rep, 2024. 14(1): p. 178.

15. Melamud, M.M., et al., NETosis-Related Biomarkers in Systemic Lupus Erythematosus, Rheumatoid Arthritis, Psoriatic Arthritis and Ankylosing Spondylitis: A Comparative Study. Int J Mol Sci, 2025. 26(24).

16. Jin, K., et al., Differences in cognitive impairment and its correlation with circulating cell-free mitochondrial DNA in medication-free depression and bipolar depression patients. J Affect Disord, 2025. 369: p. 765–771.

17. Lindqvist, D., et al., Circulating cell-free mitochondrial DNA, but not leukocyte mitochondrial DNA copy number, is elevated in major depressive disorder. Neuropsychopharmacology, 2018. 43(7): p. 1557–1564.

18. Husakova, M., et al., Mental health in patients with rheumatoid arthritis and axial spondyloarthritis: a cross-sectional, case-control tertiary centre study from Czechia. BMJ Open, 2025. 15(12): p. e103686.

19. Rabin, R. and F. de Charro, EQ-5D: a measure of health status from the EuroQol Group. Ann Med, 2001. 33(5): p. 337–43.

20. Rudwaleit, M., et al., The development of Assessment of SpondyloArthritis international Society classification criteria for axial spondyloarthritis (part II): validation and final selection. Ann Rheum Dis, 2009. 68(6): p. 777–83.

21. Sieper, J., et al., The Assessment of SpondyloArthritis international Society (ASAS) handbook: a guide to assess spondyloarthritis. Ann Rheum Dis, 2009. 68 Suppl 2: p. ii1–44.

22. Sheehan, D.V., et al., The Mini-International Neuropsychiatric Interview (M.I.N.I.): the development and validation of a structured diagnostic psychiatric interview for DSM-IV and ICD-10. J Clin Psychiatry, 1998. 59 Suppl 20: p. 22–33;quiz 34-57.

23. Pettersson, A., et al., The Mini-International Neuropsychiatric Interview is useful and well accepted as part of the clinical assessment for depression and anxiety in primary care: a mixed-methods study. BMC Fam Pract, 2018. 19(1): p. 19.

24. Beck, A.T., et al., An inventory for measuring clinical anxiety: psychometric properties. J Consult Clin Psychol, 1988. 56(6): p. 893–7.

25. Beck, A.T., R.A. Steer, and M.G. Carbin, Psychometric properties of the Beck Depression Inventory: Twenty-five years of evaluation. Clinical Psychology Review, 1988. 8(1): p. 77–100.

26. Julian, L.J., Measures of anxiety: State-Trait Anxiety Inventory (STAI), Beck Anxiety Inventory (BAI), and Hospital Anxiety and Depression Scale-Anxiety (HADS-A). Arthritis Care Res (Hoboken), 2011. 63 Suppl 11(0 11): p. S467–72.

27. Ahmad, S., et al., Simultaneous extraction of nuclear and mitochondrial DNA from human blood. Genes Genet Syst, 2007. 82(5): p. 429–32.

28. Brynychova, I., et al., Immunoregulatory properties of cell-free DNA in plasma of celiac disease patients - A pilot study. Autoimmunity, 2019. 52(2): p. 88–94.

29. Lindqvist, D., et al., Increased plasma levels of circulating cell-free mitochondrial DNA in suicide attempters: associations with HPA-axis hyperactivity. Transl Psychiatry, 2016. 6(12): p. e971.

30. Cawthon, R.M., Telomere length measurement by a novel monochrome multiplex quantitative PCR method. Nucleic Acids Res, 2009. 37(3): p. e21.

31. Salem, R., et al., DNA integrity index as a potential molecular biomarker in colorectal cancer. Egyptian Journal of Medical Human Genetics, 2020. 21(1): p. 38.

32. Salk, R.H., J.S. Hyde, and L.Y. Abramson, Gender differences in depression in representative national samples: Meta-analyses of diagnoses and symptoms. Psychol Bull, 2017. 143(8): p. 783–822.

33. Mac Giollabhui, N., et al., Role of inflammation in depressive and anxiety disorders, affect, and cognition: genetic and non-genetic findings in the lifelines cohort study. Transl Psychiatry, 2025. 15(1): p. 164.

34. Verhoeven, J.E., et al., Depression, telomeres and mitochondrial DNA: between- and within-person associations from a 10-year longitudinal study. Mol Psychiatry, 2018. 23(4): p. 850–857.

35. Park, S.S., H. Jeong, and A.C. Andreazza, Circulating cell-free mitochondrial DNA in brain health and disease: A systematic review and meta-analysis. World J Biol Psychiatry, 2022. 23(2): p. 87–102.

36. Chattopadhyay, A., H. Tak, and B.H. Naick, Evaluating cell-free circulatory mitochondrial DNA as a comprehensive biomarker for stress: meta-analysis of psychological and physiological stress responses. Biomarkers, 2025. 30(4): p. 295–303.

37. Kageyama, Y., et al., The relationship between circulating mitochondrial DNA and inflammatory cytokines in patients with major depression. J Affect Disord, 2018. 233: p. 15–20.

38. Osimo, E.F., et al., Inflammatory markers in depression: A meta-analysis of mean differences and variability in 5,166 patients and 5,083 controls. Brain Behav Immun, 2020. 87: p. 901–909.

39. Mendes-Silva, A.P., et al., Decreased Mitochondrial DNA Integrity and Elevated Inflammatory Markers in Late-Life Depression: A Longitudinal Study. Biol Psychiatry Glob Open Sci, 2025. 5(6): p. 100598.

40. Xia, P., et al., Simultaneous quantitative assessment of circulating cell-free mitochondrial and nuclear DNA by multiplex real-time PCR. Genet Mol Biol, 2009. 32(1): p. 20–4.

41. Qian, H., et al., Associations of Platelet Count with Inflammation and Response to Anti-TNF-α Therapy in Patients with Ankylosing Spondylitis. Front Pharmacol, 2020. 11: p. 559593.

42. Umetani, N., et al., Increased integrity of free circulating DNA in sera of patients with colorectal or periampullary cancer: direct quantitative PCR for ALU repeats. Clin Chem, 2006. 52(6): p. 1062–9.

43. Blaschke, F., et al., C-reactive protein induces apoptosis in human coronary vascular smooth muscle cells. Circulation, 2004. 110(5): p. 579–87.

44. Gershov, D., et al., C-Reactive protein binds to apoptotic cells, protects the cells from assembly of the terminal complement components, and sustains an antiinflammatory innate immune response: implications for systemic autoimmunity. J Exp Med, 2000. 192(9): p. 1353–64.

45. Liu, W., et al., The association between C-reactive protein-to-lymphocyte ratio and telomere length: A cross-sectional population-based study. Medicine (Baltimore), 2025. 104(41): p. e45106.

46. Gardner, J.P., et al., Rise in insulin resistance is associated with escalated telomere attrition. Circulation, 2005. 111(17): p. 2171–7.

47. Bekaert, S., et al., Telomere length and cardiovascular risk factors in a middle-aged population free of overt cardiovascular disease. Aging Cell, 2007. 6(5): p. 639–47.

48. Mistegaard, C., et al., Complement system activation is associated with spinal radiographic progression in axial spondyloarthritis after 2 years of follow-up: findings from the CONSUL RCT. RMD Open, 2025. 11(4).

49. Kyo, S., et al., Estrogen activates telomerase. Cancer Res, 1999. 59(23): p. 5917–21.

50. Rusman, T., R.E. van Bentum, and I.E. van der Horst-Bruinsma, Sex and gender differences in axial spondyloarthritis: myths and truths. Rheumatology (Oxford), 2020. 59(Suppl4): p. iv38–iv46.

