## Supplementary Material Tables for "Cell-free telomeric DNA and integrity indices in axial spondyloarthritis: associations with somatic and mental burden — a case-control study"

**Supplementary Table 1 Demographic and clinical characteristics across sexes**

| **Feature** | **axSpA males (n=51)** | **axSpA females (n=28)** | **HC males (n=62)** | **HC females (n=26)** |
| --- | --- | --- | --- | --- |
| **Age, median (IQR)** | 41.00 (33.50 - 48.00) | 42.00 (38.00 - 48.25) | 37.00 (25.00 - 47.00)^***^ | 47.00 (43.75 - 52.75) |
| **BMI, median (IQR)** | 25.50 (23.77 - 28.74) | 25.06 (22.30 - 30.54) | 26.66 (24.17 - 29.65) | 26.80 (21.38 - 30.22) |
| **Marital status** – **1/2, %** | 35.29/64.71 | 28.57/71.43 | 51.61/48.39 | 30.77/69.23 |
| **Adults in household** – **=1/ >1, %** | 19.61/80.39 | 28.57/71.43 | 24.19/75.81 | 30.77/69.23 |
| **City population** – **<20k/20-100/>100k, %** | 49.02/13.73/37.25 | 42.86/17.86/3929 | 32.26/9.68/58.06 | 38.46/11.54/50.00 |
| **Work status** – **1/2/3, %** | 92.20/3.90/3.90^**^ | 75.00/21.40/3.57 | 77.40/1.60/21.00 | 88.50/11.50/0.00 |
| **HAQ median (IQR)** | 0.12 (0.00 - 0.50)^***^ | 0.25 (0.00 - 0.75)^**^ | 0.00 (0.00 - 0.00) | 0.00 (0.00 - 0.25) |
| **5D-EQ, median (IQR)** | 0.80 (0.69 - 0.93)^****^ | 0.78 (0.68 - 0.80)^***^ | 1.00 (1.00 - 1.00) | 1.00 (0.80 - 1.00) |
| **Comorbidities** – **untracked dis./up to 2/>2, %** | 58.82/37.25/3.92 | 46.43/46.43/7.14 | 67.74/29.03/3.23^**^ | 34.62/57.69/7.69 |
| **Alcohol consumption below/in line/above WHO recommendations^a^, %** | 82.35/13.73/3.92 | 89.29/10.71/0.00 | 80.65/11.29/8.06 | 80.77/19.23/0.00 |
| **Meets WHO physical activity recommendations^b^, %** | 80.39 ^**^ | 46.43^**^ | 70.97 | 50.00 |
| **Disease duration since first symtom, median (IQR)** | 12.26 (6.36 - 20.22) | 11.04 (6.96 - 14.57) |  |  |
| **Disease duration since diagnosis, median (IQR)** | 5.91 (2.84 - 10.29) | 6.63 (3.35 - 9.63) |  |  |
| **HLA-B27 positive, %** | 94.12 | 89.29 |  |  |
| **Non-radiographic/Radiographic axSpA, %** | 27.45/72.55^**^ | 60.71/39.29^**^ |  |  |
| **AxSpA restricted to SIJ/Spine involvement, %** | 54.90/45.10^***^ | 89.30/10.70^***^ |  |  |
| **Hip involvement, %** | 35.29^**^ | 10.71^**^ |  |  |
| **Current peripheral arthritis/enthesitis/dactylitis, %** | 13.73/13.73/3.9 | 7.14/25.00/3.57 |  |  |
| **CRP, median (IQR)** | 2.50 (1.27 - 6.57) | 1.98 (0.55 - 8.12) |  |  |
| **ASDAS-CRP, median (IQR)** | 1.74 (1.05 - 2.76) | 1.98 (1.24 - 2.76) |  |  |
| **ASDAS-CRP, without activity (0-1.3), %** | 31.37 | 28.57 |  |  |
| **ASDAS-CRP, mild activity (1.4-2.0), %** | 35.29 | 28.57 |  |  |
| **ASDAS-CRP, moderate activity (2.1-3.4), %** | 21.57 | 32.14 |  |  |
| **ASDAS-CRP, severe activity (>3.5), %** | 11.76 | 10.71 |  |  |
| **BASDAI, median (IQR)** | 2.80 (1.10 - 4.10) | 2.85 (1.75 - 5.30) |  |  |
| **BASFI, median (IQR)** | 0.90 (0.20 - 2.70) | 1.65 (0.18 - 3.10) |  |  |
| **Medication** – **bDMARDs, %** | 27.45^*^ | 3.57^*^ |  |  |
| **Medication** – **csDMARDs, %** | 7.84 | 10.71 |  |  |

Abbreviations: axSpA – Axial spondyloarthritis; HC – Healthy controls; BMI – Body mass index; Marital status: 1 – married/partner, 2 – single/divorced/widowed; WHO – World health organisation; HAQ – Health assessment questionnaire; 5D-EQ – Five dimensional quality of life; Work status: 1 – full-time/part-time; 2 – sick-leave/unemployed/diseased pension; 3 – student/parental-leave/pension; IBP – Inflammatory back pain, CRP – C reactive protein, ASDAS-CRP – Ankylosing spondylitis (AS) disease activity score with CRP; BASDAI – Bath AS disease activity index; BASFI – Bath AS functional index; BASMI – Bath AS Mobility index; SIJ – Sacroiliac joint; DMARDs – Disease modifying drugs; b – Biologic; cs – conventional syntetic.

^a^ WHO Recommendation for alcohol consumption: 2 standard units for women and 3 standard units for men; standard units: hard alcohol – 0.04l shot 1.3 units, beer 10° 0.33l 1 unit, beer 12° 1.5 units, wine 1dcl 1 unit.

^b^ WHO Physical activity: 30 minutes of moderate-intensity physical activity 5 times a week or 20 minutes of high-intensity physical activity 3 times a week.

Different sets of comparisons are indicated by color-coded symbols: axSpA males vs axSpA females; axSpA males vs. HC males; axSpA females vs. HC females; HC males vs. HC females

Statistical signifikance (uncorrected): * p<0.05, ** p<0.01, *** p<0.001, **** p<0.0001; Bonferroni correction was applied separately within each set of comparisons (2 comparisons per set; adjusted α = 0.025).

**Supplementary Table 2 Mental burden in patients with axSpA and their healthy counterparts**

| **Feature** | **axSpA (n=79)** | **HC (n=88)** | **HC males (n=62)** | **HC females (n=26)** | **axSpA males (n=51)** | **axSpA females (n=28)** |
| --- | --- | --- | --- | --- | --- | --- |
| **Presence/absence of any MD, M.I.N.I., %** | 32.91/67.09**** | 6.82/93.18 | 9.68/90.32* | 0.00/100.00 | 27.45/72.55* | 42.86/57.14*** |
| **Presence/absence AffectD, M.I.N.I., %** | 26.58/73.42**** | 2.27/97.73 | 3.23/96.77 | 0.00/100.00 | 21.57/78.43** | 35.71/64.29*** |
| **Presence/absence AnxD, M.I.N.I., %** | 13.92/86.08* | 4.55/95.45 | 6.45/93.55 | 0.00/100.00 | 11.76/88.24 | 17.86/82.14* |
| **Depression/without depression BDI-II, %** | 20.25/79.75*** | 2.27/97.73 | 1.61/98.39 | 3.85/96.15 | 19.61/80.39** | 21.43/78.57 |
| **BDI-II, mean (SD)** | 6.73 (± 6.91)**** | 2.84 (± 3.47) | 2.55 (± 3.24) | 3.54 (± 3.93) | 6.08 (± 6.07)*** | 7.93 (± 8.22)* |
| **BDI-II – without/mild/moderate/ severe depression, %** | 79.75/15.19/3.80/1.27*** | 97.73/2.27/0.00/0.00 | 98.39/1.61/0.00/0.00 | 96.15/3.85/0.00/0.00 | 80.39/17.65/1.96/0.00** | 78.57/10.71/7.14/3.57* |
| **BDI-II Somatic, mean (SD)** | 3.65 (± 3.46)**** | 1.64 (± 2.20) | 1.29 (± 1.79) | 2.46 (± 2.83) | 3.27 (± 3.24)*** | 4.32 (± 3.79)* |
| **BDI-II Cognitive, mean (SD)** | 1.48 (± 2.44)** | 0.57 (± 1.19) | 0.61 (± 1.32) | 0.46 (± 0.81) | 1.31 (± 2.11)* | 1.79 (± 2.96)* |
| **BDI-II Affective, mean (SD)** | 1.61 (± 1.84)**** | 0.64 (± 1.10) | 0.65 (± 1.13) | 0.62 (± 1.02) | 1.49 (± 1.60)** | 1.82 (± 2.21)* |
| **Anxiety/without anxiety BAI, %** | 22.78/77.22 | 11.36/88.64 | 8.06/91.94 | 19.23/80.77 | 15.69/84.31 | 35.71/64.29 |
| **BAI, mean (SD)** | 6.75 (± 8.65)** | 3.89 (± 3.92) | 3.37 (± 3.52) | 5.12 (± 4.57) | 5.02 (± 6.16)* | 9.89 (± 11.41)* |
| **BAI – without/mild/moderate/ severe anxiety, %** | 77.22/16.46/3.80/2.53* | 88.64/11.36/0.00/0.00 | 91.94/8.06/0.00/0.00 | 80.77/19.23/0.00/0.00 | 84.31/13.73/0.00/1.96 | 64.29/21.43/10.71/3.57 |

Abbreviations: axSpA – Axial spondyloarthritis; HC – Healthy controls; MD – Mental disorder according to M.I.N.I. (Mini-International Neuropsychiatric Interview); BDI-II – Beck Depression Inventory: without depression (0-13), mild (14-19), moderate (20-28), severe (29-63); BAI – Beck Anxiety Inventory: without anxiety (0-9), mild (10-18), moderate (19-29), severe (30-63).

Different sets of comparisons are indicated by color-coded symbols: axSpA vs. HC; axSpA males vs. axSpA females; axSpA males vs. HC males; axSpA females vs. HC females; HC males vs. HC females

Statistical significance: *p<0.05; ** p<0.01; *** p<0.001; **** p<0.0001; Bonferroni correction was applied separately within each set of comparisons: for sex-stratified comparisons and within-group sex comparisons (2 comparisons per set; adjusted α = 0.025). No correction was applied for the overall comparison (single test).

**Supplementary Table 3 Variability of the mental burden among axSpA patients**

| **Feature** | **r-axSpA (n=48)** | **nr-axSpA (n=31)** | **axSpA SIJ limited (n=53)** | **axSpA Spine involvement (n=26)** | **ASDAS-CRP ≥ 1.4 (n=50)** | **ASDAS-CRP < 1.4 (n=29)** |
| --- | --- | --- | --- | --- | --- | --- |
| **Presence/absence of any MD, M.I.N.I., %** | 35.42/64.58 | 29.03/70.97 | 33.96/66.04 | 30.77/69.23 | 44.00/56.00** | 13.79 /86.21 |
| **Depression/without depression BDI-II, %** | 25.00/75.00 | 12.90/87.10 | 15.09/84.91 | 30.77/69.23 | 30.00/70.00*** | 3.45/96.55 |
| **BDI-II, mean (SD)** | 6.00 (3.00 - 12.50) | 3.00 (0.00 - 5.00) | 5.85 (± 7.05) | 8.54 (± 6.38) | 6.50 (4.00 - 14.00)**** | 3.00 (0.00 - 4.00) |
| **BDI-II – without/mild/moderate/ severe depression, %** | 75.00/20.83/4.17/0.00 | 87.10/6.45/3.23/3.23 | 84.91/9.43/3.77/1.89 | 69.23/26.92/3.85/0.00 | 70.00/22.00/6.00/2.00*** | 96.55/3.45/0.00/0.00 |
| **BDI-II Somatic, mean (SD)** | 4.17 (± 3.50) | 2.84 (± 3.30) | 3.08 (± 3.32)* | 4.81 (± 3.52) | 4.86 (± 3.54)**** | 1.55 (± 2.06) |
| **BDI-II Cognitive, mean (SD)** | 1.58 (± 2.18) | 1.32 (± 2.82) | 1.43 (± 2.52) | 1.58 (± 2.32) | 1.96 (± 2.80)** | 0.66 (± 1.32) |
| **BDI-II Affective, mean (SD)** | 1.98 (± 1.68)* | 1.03 (± 1.94) | 1.34 (± 1.86) | 2.15 (± 1.69) | 2.16 (± 1.99)**** | 0.66 (± 0.97) |
| **Anxiety/without anxiety BAI, %** | 22.92/77.08 | 22.58/77.42 | 22.64/77.36 | 23.08/76.92 | 34.00/66.00*** | 3.45/96.55 |
| **BAI, median (IQR)** | 5.00 (2.00 - 9.00) | 2.00 (1.00 - 6.00) | 5.64 (± 6.90) | 9.00 (± 11.25) | 6.00 (2.00 - 13.00)*** | 2.00 (1.00 - 4.00) |
| **BAI – without/mild/moderate/ severe anxiety, %** | 77.08/16.67/2.08/4.17 | 77.42/16.13/6.45/0.00 | 77.36/16.98/5.66/0.00 | 76.92/15.38/0.00/7.69 | 66.00/24.00/6.00/4.00*** | 96.55/3.45/0.00/0.00 |

Abbreviations: axSpA – Axial spondyloarthritis; r-radiographic; nr-nonradiographic; MD – Mental disorder according to M.I.N.I. (Mini-International Neuropsychiatric Interview); BDI-II – Beck depression inventory: without depression (0-13), mild (14-19), moderate (20-28), severe (29-63); BAI – Beck Anxiety Inventory: without anxiety (0-9), mild (10-18), moderate (19-29), severe (30-63).

Different sets of comparisons are indicated by color-coded symbols: r-axSpA vs. nr-axSpA; axSpA SIJ limited vs. axSpA Spine involvement; axSpA ASDAS ≥ 1.4 vs. axSpA ASDAS < 1.4

Statistical significance (uncorrected) * p<0.05; ** p<0.01; *** p<0.001; **** p<0.0001; Bonferroni correction was applied for multiple comparisons within each variable (3 comparisons; adjusted α = 0.0167); therefore, only results with p < 0.0167 were considered statistically significant.

**Supplementary Table 4 Plasma levels of the circulating DNA fragments in axSpA and HC across sexes**

| **Feature** | **HC males (n=62)** | **HC females (n=26)** | **axSpA males (n=51)** | **axSpA females (n=28)** |
| --- | --- | --- | --- | --- |
| **cf-nDNA (ng/ml), median (IQR)** | 22.25 (14.67 - 35.84) | 16.03 (10.25 - 20.63) | 17.67 (11.87 - 24.74) | 14.88 (10.64 - 23.71) |
| **cf-mtDNA (ng/ml), median (IQR)** | 1.76 (1.20 - 2.53) | 1.34 (0.64 - 2.54) | 2.12 (1.09 - 4.19) | 2.50 (1.28 - 4.72)* |
| **DII, median (IQR)** | 1.18 (0.94 - 1.38)* | 1.33 (1.18 - 1.54) | 0.90 (0.78 - 1.15)**^/^* | 1.34 (1.09 - 1.46) |
| **T/S ratio, median (IQR)** | 62.63 (38.15 - 124.09) | 105.52 (56.09 - 153.57) | 53.30 (38.68 - 82.13)*** | 125.71 (86.23 - 243.04) |

Abbreviations: axSpA – Axial spondyloarthritis; HC – Healthy controls; cf-n/mtDNA – Cell-free circulating nuclear/mitochondrial DNA; DII – DNA integrity index; T/S – Telomere-to-single copy gene; IQR – Interquartile range.

Different comparisons are indicated by color-coded symbols: axSpA males vs. axSpA females; axSpA males vs. HC males; axSpA females vs. HC females; HC males vs. HC females

Statistical significance: *p < 0.05; **p < 0.01; ***p < 0.001; P-values are uncorrected; no adjustment for multiple testing was applied due to the exploratory nature of the analyses.

**Supplementary Table 5 Spearman's correlation of DNA fragments and clinical variables in HC**

|  | **cf-nDNA** | **cf-mtDNA** | **DII** | **T/S ratio** |
| --- | --- | --- | --- | --- |
| **cf-nDNA** | 1 | ***0,395 | -0,071 | ***-0,780 |
| **cf-mtDNA** | ***0,395 | 1 | -0,133 | *-0,232 |
| **DII** | -0,071 | -0,133 | 1 | 0,075 |
| **T/S ratio** | ***-0,780 | *-0,232 | 0,075 | 1 |
| **BMI** | 0,132 | 0,024 | **0,299 | -0,127 |
| **Comorbidities** | 0,013 | 0,142 | 0,173 | *-0,237 |
| **WHO Physical activity** | -0,025 | 0,063 | -0,064 | 0,031 |
| **WHO Alcohol consumption** | 0,103 | 0,105 | -0,042 | -0,141 |
| **HAQ** | -0,187 | 0,023 | 0,018 | 0,050 |
| **5D-EQ** | 0,072 | 0,062 | 0,069 | 0,092 |
| **MD (M.I.N.I.)** | -0,006 | *0,113 | 0,002 | -0,012 |
| **BDI-II** | -0,006 | -0,045 | -0,087 | -0,157 |
| **BAI** | -0,033 | -0,075 | 0,069 | -0,075 |
| **BDI-II Somatic Score** | 0,009 | -0,041 | -0,101 | -0,161 |
| **BDI-II Cognitive Score** | -0,051 | -0,011 | -0,077 | -0,144 |
| **BDI-II Affective Score** | -0,141 | -0,199 | 0,004 | -0,030 |
| **Male sex** | *0,255 | 0,111 | *-0,229 | -0,163 |
| **Age** | -0,035 | -0,021 | 0,137 | -0,142 |

Abbreviations: HC – Healthy controls; cf-n/mtDNA – Cell-free circulating nuclear/mitochondrial DNA; DII – DNA integrity index; T/S – Telomere-to-single copy gene; BMI – Body mass index; WHO – World health organization; HAQ – Health assessment questionnaire; 5D-EQ – Five dimensional quality of life; MD – Mental disorder according to M.I.N.I. (Mini-International Neuropsychiatric Interview); BDI-II – Beck Depression Inventory; BAI – Beck Anxiety Inventory.

Statistical significance: *<0.05; **<0.01; ***<0.001

Due to the exploratory nature of the biomarker analyses, no adjustment for multiple testing was applied. Reported p-values are uncorrected and should be interpreted with caution as hypothesis-generating findings.

**Supplementary Table S6 Spearman's correlation of DNA fragments and clinical variables in HC across sexes**

|  | **Males** | | | | **Females** | | | |
| --- | --- | --- | --- | --- | --- | --- | --- | --- |
|  | **cf-nDNA** | **cf-mtDNA** | **DII** | **T/S ratio** | **cf-nDNA** | **cf-mtDNA** | **DII** | **T/S ratio** |
| **cf-nDNA** | 1 | *0,298 | 0,054 | -0,758 | 1 | ***0,618 | -0,115 | ***  -0,802 |
| **cf-mtDNA** | *0,298 | 1 | -0,082 | -0,047 | ***0,618 | 1 | -0,144 | ***  -0,627 |
| **DII** | 0,054 | -0,082 | 1 | -0,074 | -0,115 | -0,144 | 1 | 0,288 |
| **T/S ratio** | -0,757 | -0,047 | -0,074 | 1 | ***-0,802 | ***-0,627 | 0,288 | 1 |
| **BMI** | 0,0105 | -0,009 | **0,361 | -0,132 | 0,249 | -0,001 | 0,235 | 0,001 |
| **Comorbidities** | 0,034 | 0,111 | *0,266 | *-0,261 | 0,315 | 0,329 | -0,202 | *-0,420 |
| **WHO Physical activity** | -0,143 | -0,030 | 0,008 | 0,167 | 0,015 | 0,149 | -0,067 | -0,159 |
| **WHO Alcohol consumption** | 0,093 | 0,051 | 0,005 | -0,149 | 0,137 | 0,176 | -0,084 | -0,176 |
| **HAQ** | -0,047 | 0,034 | -0,045 | -0,114 | -0,210 | 0,086 | -0,019 | 0,116 |
| **5D-EQ** | -0,004 | 0,036 | 0,176 | 0,230 | 0,172 | 0,129 | -0,095 | -0,087 |
| **MD (M.I.N.I.)** | -0,069 | 0,120 | 0,044 | 0,024 | NA | NA | NA | NA |
| **BDI-II** | 0,080 | -0,054 | -0,049 | *-0,280 | -0,145 | -0,047 | -0,197 | 0,060 |
| **BAI** | -0,029 | -0,118 | 0,155 | -0,030 | 0,098 | 0,054 | -0,252 | -0,263 |
| **BDI-II Somatic Score** | 0,112 | -0,050 | -0,064 | *-0,316 | -0,045 | 0,035 | -0,261 | 0,021 |
| **BDI-II Cognitive Score** | -0,034 | 0,020 | -0,038 | -0,183 | -0,162 | -0,123 | -0,100 | -0,050 |
| **BDI-II Affective Score** | -0,093 | *-0,283 | 0,020 | -0,109 | -0,373 | -0,128 | -0,033 | 0,151 |
| **Age** | 0,007 | -0,044 | 0,131 | *-0,251 | 0,255 | 0,200 | -0,131 | -0,147 |

Abbreviations: HC – Healthy controls; cf-n/mtDNA – Cell-free circulating nuclear/mitochondrial DNA; DII – DNA integrity index; T/S – Telomere-to-single copy gene; BMI – Body mass index; WHO – World health organization; HAQ – Health assessment questionnaire; 5D-EQ – Five dimensional quality of life; MD – Mental disorder according to M.I.N.I. (Mini-International Neuropsychiatric Interview); BDI-II – Beck Depression Inventory; BAI – Beck Anxiety Inventory.

Statistical significance: * p<0.05; ** p<0.01; *** p<0.001. Due to the exploratory nature of the biomarker analyses, no adjustment for multiple testing was applied. Reported p-values are uncorrected and should be interpreted with caution as hypothesis-generating findings.

**Supplementary Table S7 Spearman's correlation of DNA fragments and clinical variables in axSpA patients across sexes**

|  | **Males** | | | | **Females** | | | |
| --- | --- | --- | --- | --- | --- | --- | --- | --- |
|  | **cf-nDNA** | **cf-mtDNA** | **DII** | **T/S ratio** | **cf-nDNA** | **cf-mtDNA** | **DII** | **T/S ratio** |
| **cf-nDNA** | 1 | **0,441 | -0,118 | ***-0,622 | 1 | *0,398 | 0,094 | **-0,513 |
| **cf-mtDNA** | **0,441 | 1 | -0,160 | -0,170 | *0,398 | 1 | *-0,413 | -0,256 |
| **DII** | -0,118 | -0,160 | 1 | *0,328 | 0,094 | *-0,414 | 1 | 0,290 |
| **T/S ratio** | ***-0,622 | -0,170 | *0,328 | 1 | **-0,513 | -0,256 | 0,290 | 1 |
| **BMI** | 0,231 | -0,018 | -0,154 | -0,241 | 0,328 | 0,124 | 0,035 | -0,374 |
| **Comorbidities** | *0,282 | 0,203 | -0,063 | **-0,416 | -0,156 | -0,227 | 0,121 | 0,048 |
| **WHO Physical activity** | 0,059 | -0,037 | -0,065 | -0,074 | -0,199 | -0,297 | -0,106 | -0,235 |
| **WHO Alcohol consumption** | -0,041 | -0,041 | 0,092 | -0,108 | -0,021 | 0,179 | 0,136 | -0,007 |
| **HAQ** | 0,244 | 0,111 | *-0,292 | *-0,283 | 0,130 | -0,038 | 0,088 | -0,111 |
| **5D-EQ** | -0,184 | 0,019 | 0,056 | *0,290 | 0,186 | 0,158 | -0,091 | -0,022 |
| **MD (M.I.N.I.)** | -0,110 | -0,078 | -0,094 | -0,064 | -0,286 | -0,170 | 0,071 | 0,366 |
| **BDI-II** | 0,106 | -0,035 | -0,097 | -0,253 | -0,035 | -0,021 | -0,081 | -0,086 |
| **BAI** | 0,187 | -0,041 | 0,025 | -0,212 | -0,131 | -0,231 | 0,118 | 0,037 |
| **BDI-II Somatic scores** | 0,064 | -0,004 | -0,050 | -0,231 | -0,015 | 0,022 | -0,020 | -0,081 |
| **BDI-II Cognitive scores** | 0,021 | 0,053 | -0,102 | -0,114 | -0,258 | -0,197 | -0,047 | 0,084 |
| **BDI-II Affective scores** | 0,102 | -0,216 | -0,091 | -0,199 | 0,047 | -0,053 | -0,023 | -0,112 |
| **DisDur since symptoms** | 0,051 | -0,053 | -0,086 | -0,224 | 0,062 | -0,269 | 0,123 | -0,001 |
| **DisDur since diagnosis** | 0,004 | -0,034 | 0,039 | -0,123 | 0,368 | 0,021 | 0,180 | -0,028 |
| **Radiographic axSpA** | 0,044 | -0,202 | -0,115 | -0,120 | 0,218 | 0,055 | -0,244 | -0,262 |
| **Spine involvement** | 0,044 | -0,133 | -0,205 | -0,148 | 0,365 | -0,064 | 0,107 | -0,136 |
| **CRP** | 0,191 | 0,012 | **-0,383 | *-0,289 | 0,140 | 0,015 | -0,137 | **-0,527 |
| **ASDAS-CRP** | 0,066 | -0,093 | -0,196 | -0,238 | -0,080 | 0,003 | 0,015 | -0,365 |
| **BASDAI** | 0,042 | -0,004 | -0,025 | -0,200 | -0,176 | -0,081 | 0,053 | -0,155 |
| **BASFI** | 0,223 | 0,083 | -0,263 | *-0,409 | 0,084 | 0,026 | 0,113 | -0,143 |
| **Medication bDMARDs** | -0,077 | -0,094 | 0,224 | 0,044 | -0,416 | *0,01 | -0,280 | 0,132 |
| **HLA-B27 positivity** | 0,082 | 0 | 0,076 | 0,057 | 0,164 | 0,150 | 0,029 | -0,193 |
| **Age** | 0,115 | 0,073 | -0,147 | *-0,328 | 0,287 | 0,127 | 0,073 | -0,125 |

Abbreviations: axSpA – Axial spondyloarthritis; cf-n/mtDNA – Cell-free circulating nuclear/mitochondrial DNA; DII – DNA integrity index; T/S – Telomere-to-single copy gene; BMI – Body mass index; WHO – World health organization; HAQ – Health assessment questionnaire; 5D-EQ – Five dimensional quality of life; MD – Mental disorder according to M.I.N.I. (Mini-International Neuropsychiatric Interview); BDI-II – Beck Depression Inventory; BAI – Beck Anxiety Inventory; DisDur – Disease duration; CRP – C reactive protein; ASDAS-CRP – Ankylosing spondylitis (AS) disease activity score with CRP; BASDAI – Bath AS disease activity index; BASFI – Bath AS functional index; DMARDs – Disease modifying drugs; b – Biologic; HLA – Human leukocyte antigen.

Statistical significance: * p<0.05; ** p<0.01; *** p<0.001

**Supplementary Material Table S8 Linear regression models for mental health, disease activity and cf-DNA in axSpA**

| **BAI** |  |  |  |  |  | |  | |
| --- | --- | --- | --- | --- | --- | --- | --- | --- |
| Age | **coef.** | **SE** | **t** | **p** | **0.025** | | **0.975** | |
|  | 0.1190 | 0.020 | 5.969 | <0.0001 | 0.079 | | 0.159 | |
| Male sex | -5.1330 | 0.595 | -8.624 | <0.0001 | -6.320 | | -3.946 | |
| cf-nDNA | -0.0317 | 0.016 | -2.047 | <0.05 | -0.063 | | -0.001 | |
| cf-mtDNA | -0.2459 | 0.070 | -3.490 | <0.001 | -0.386 | | -0.105 | |
| DII | 1.3974 | 0.447 | 3.125 | <0.01 | 0.506 | | 2.289 | |
| T/S ratio | -0.0032 | 0.002 | -1.640 | 0.105 | -0.007 | | 0.001 | |
| CRP | 4.0110 | 0.342 | 11.733 | <0.0001 | 3.329 | | 4.693 | |
| Intercept | -1.7789 | 1.504 | -1.183 | 0.241 | -4.778 | | 1.220 | |
| **BDI-II** |  |  |  |  |  | |  | |
|  | **coef.** | **SE** | **t** | **p** | **0.025** | | **0.975** | |
| Age | 0.0144 | 0.019 | 0.740 | 0.462 | -0.024 | | 0.053 | |
| Male sex | -1.1819 | 0.481 | -2.456 | <0.05 | -2.142 | | -0.222 | |
| cf-nDNA | -0.0267 | 0.017 | -1.558 | 0.124 | -0.061 | | 0.007 | |
| cf-mtDNA | -0.0562 | 0.051 | -1.090 | 0.279 | -0.159 | | 0.047 | |
| DII | 1.0316 | 0.535 | 1.928 | 0.058 | -0.035 | | 2.099 | |
| T/S ratio | 0.0030 | 0.002 | 1.316 | 0.192 | -0.002 | | 0.008 | |
| CRP | 4.2578 | 0.333 | 12.794 | <0.001 | 3.594 | | 4.921 | |
| Intercept | 1.4366 | 1.473 | -0.975 | 0.333 | -4.374 | | 1.501 | |
| **CRP** |  |  |  |  | |  | |  |
|  | **coef.** | **SE** | **t** | **p** | | **0.025** | | **0.975** |
| Age | 0.1382 | 0.021 | 6.554 | <0.0001 | | 0.096 | | 0.180 |
| Male sex | -2.3430 | 0.483 | -4.853 | <0.0001 | | -3.305 | | -1.381 |
| cf-nDNA | 0.0508 | 0.033 | 1.550 | 0.125 | | -0.015 | | 0.116 |
| cf-mtDNA | -0.1653 | 0.077 | -2.157 | 0.034 | | -0.318 | | -0.013 |
| DII | -2.8613 | 0.544 | -5.261 | <0.0001 | | -3.946 | | -1.77 |
| T/S ratio | -0.0156 | 0.002 | -7.627 | <0.0001 | | -0.020 | | -0.012 |
| Intercept | 5.0660 | 1.086 | 4.665 | <0.0001 | | 2.901 | | 7.231 |
| **ASDAS-CRP** | |  |  |  | |  | |  |
| Age | **coef.** | **SE** | **t** | **p** | | **0.025** | | **0.975** |
|  | 0.0065 | 0.005 | 1.327 | 0.189 | | -0.003 | | 0.016 |
| Male sex | -0.0899 | 0.112 | -0.799 | 0.427 | | -0.314 | | 0.134 |
| cf-nDNA | -0.0060 | 0.003 | -2.115 | <0.05 | | -0.012 | | -0.000 |
| cf-mtDNA | 0.0025 | 0.010 | 0.266 | 0.791 | | -0.016 | | 0.022 |
| DII | 0.1735 | 0.144 | 1.209 | 0.231 | | -0.113 | | 0.460 |
| T/S ratio | 0.0004 | 0.001 | 0.686 | 0.495 | | -0.001 | | 0.002 |
| CRP | 1.2054 | 0.066 | 18.201 | <0.001 | | 1.073 | | 1.337 |
| Intercept | -0.4430 | 0.337 | -1.316 | 0.192 | | -1.114 | | 0.228 |

Abbreviations: axSpA – Axial spondyloarthritis; BAI – Beck anxiety inventory; BDI-II – Beck depression inventory-II; CRP – C reactive protein; ASDAS-CRP – Ankylosing spondylitis (AS) disease activity score with CRP; cf-n/mtDNA – Cell-free circulating nuclear/mitochondrial DNA; DII – DNA integrity index; T/S – Telomere-to-single copy gene.
